# Assisting Scalable Diagnosis Automatically via CT Images in the Combat against COVID-19

**DOI:** 10.1101/2020.05.11.20093732

**Authors:** Bohan Liu, Pan Liu, Lutao Dai, Yanlin Yang, Peng Xie, Yiqing Tan, Jicheng Du, Wei Shan, Chenghui Zhao, Qin Zhong, Xixiang Lin, Xizhou Guan, Ning Xing, Yuhui Sun, Wenjun Wang, Zhibing Zhang, Xia Fu, Yanqing Fan, Meifang Li, Na Zhang, Lin Li, Yaou Liu, Lin Xu, Jingbo Du, Zhenhua Zhao, Xuelong Hu, Weipeng Fan, Rongpin Wang, Chongchong Wu, Yongkang Nie, Liuquan Cheng, Lin Ma, Zongren Li, Qian Jia, Minchao Liu, Huayuan Guo, Gao Huang, Haipeng Shen, Weimin An, Hao Li, Jianxin Zhou, Kunlun He

## Abstract

The pandemic of coronavirus Disease 2019 (COVID-19) caused enormous loss of life globally. 1-3 Case identification is critical. The reference method is using real-time reverse transcription PCR (rRT-PCR) assays, with limitations that may curb its prompt large-scale application. COVID-19 manifests with chest computed tomography (CT) abnormalities, some even before the onset of symptoms. We tested the hypothesis that application of deep learning (DL) to the 3D CT images could help identify COVID-19 infections. Using the data from 920 COVID-19 and 1,073 non-COVID-19 pneumonia patients, we developed a modified DenseNet-264 model, COVIDNet, to classify CT images to either class. When tested on an independent set of 233 COVID-19 and 289 non-COVID-19 patients. COVIDNet achieved an accuracy rate of 94.3% and an area under the curve (AUC) of 0.98. Application of DL to CT images may improve both the efficiency and capacity of case detection and long-term surveillance.

## Main text

The world is suffering from the COVID-19 pandemic since its outbreak in December 2019.^1-3^ COVID-19 is highly contagious and infected patients can be asymptomatic but infective.^4^ As of March 29, 2020, there has been over 634,835 confirmed COVID-19 cases and 29,891 deaths worldwide.^5^ Community transmission has been increasingly reported in more than 180 countries.^5^ Improving efficiency of the current clinical pathways and the capacity of patient management are urgent needs to combat the COVID-19 during the pandemic and possible resurgence in the future.^6,7^ Case identification is a crucial first step for subsequent clinical triage and treatment optimization. The reference detection method is using the real-time reverse transcription PCR (RT-PCR) assay to detect viral RNA.^1^ A number of limitations of this assay may curb its prompt large-scale application.^8-10^

Chest computed tomography (CT) can sensitively detect manifestations of COVID-19 infections and even asymptomatic infections.^10-12^ Deep learning, an AI technology, has achieved impressive performance in analysis of CT images.^13-16^ Chest CT with the aid of deep learning offers promises to reduce the burden of prompt mass case detection, especially under the shortage of RT-PCR, and long term surveillance of this virus.^17^ We developed an automated and robust deep learning model, COVIDNet, by directly analyzing 3D CT images, to assist screening and diagnosis of COVID-19 infected patients. We provided clinical insights of the image features extracted by COVIDNet, and proposed a practical scenario on how the developed tool might improve clinical efficiency.

Two independent cohorts of 2,800 patients were retrospectively recruited for model development and external validation, including 1,430 COVID-19 patients and 1,370 non-COVID-19 patients (Figure 1). Model development used 920 COVID-19 patients and 1,073 non-COVID-19 patients, who were randomly divided into three non-overlapping sets: training, validation, and internal testing, approximately at 3:1:1 ratio (Figure 1, Supplementary Table 1, 2 and 3). External validity was evaluated using the independent external test dataset, consisting of 233 COVID-19 patients and 289 non-COVID-19 pneumonia patients (Figure 1, Supplementary Table 4 and 5).

**Figure 1.**
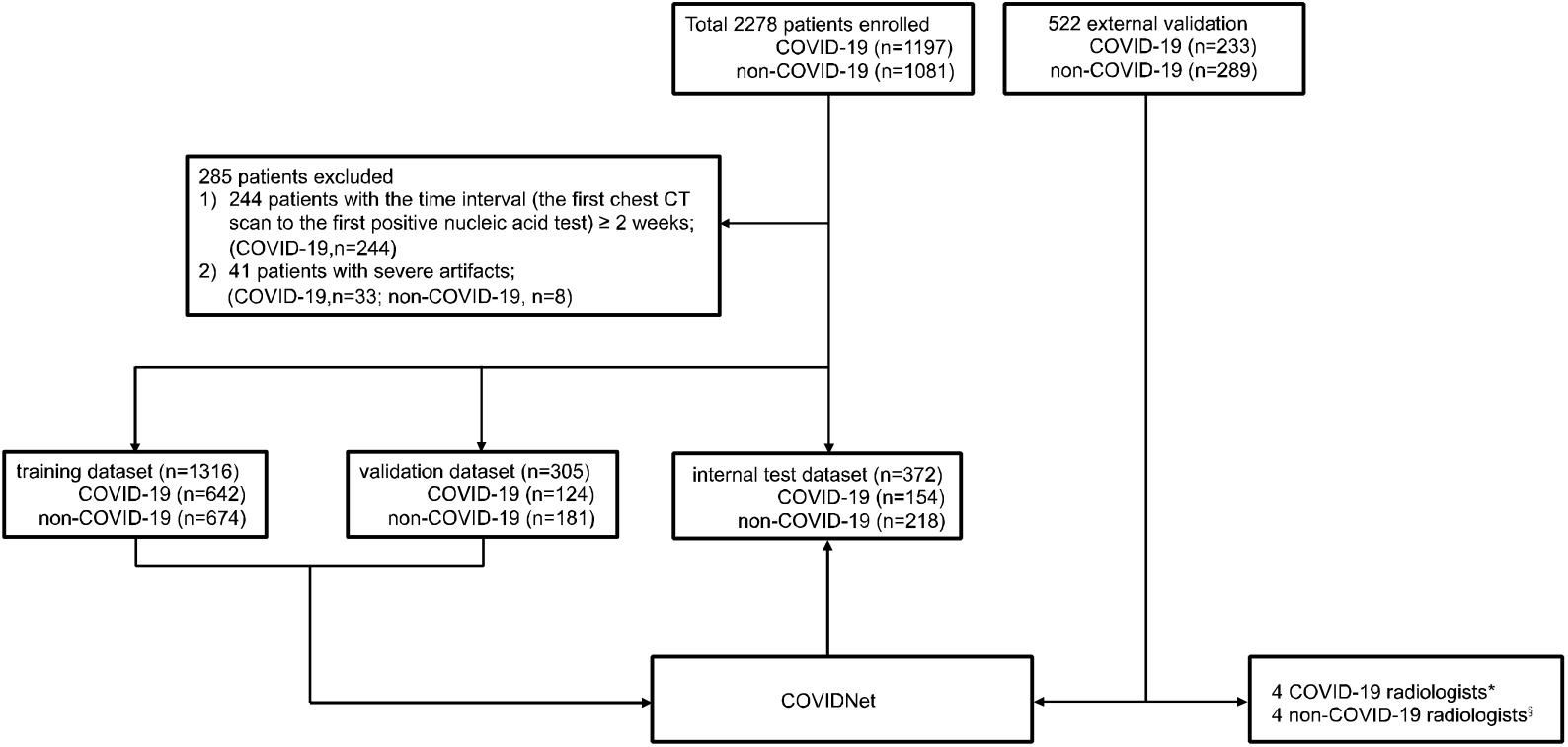
Flow diagram illustrating division of model development data into training, validation, internal test set, and external validation on the external test set. *COVID-19 radiologists: radiologists working at the COVID-19 designated hospitals; §non-COVID-19 radiologists: radiologists not working at the COVID-19 designated hospitals.

The internal test dataset included 372 patients, 41.4% (154/372) of which were confirmed COVID-19 cases. COVIDNet yielded a remarkable diagnostic performance, with an accuracy rate of 96.0% and an AUC of 0.986. More performance measures were shown in Table 1 and Extended Data Figure 1. The model performance was based on the all first CT scan of each patient. Note that our development dataset might have an overlap of 17 patients with the dataset of Li et. al,^18^ including 14 patients in the training set and 3 patients in the internal test set. Retraining and retesting after removing these patients yielded similar results (Supplementary Table 6 and Extended Data Figure 2).

**Table 1.** COVIDNet diagnostic performance on the internal test dataset. AUC=Area under the ROC curve. PPV=positive predictive value. NPV=negative predictive value.

|  | COVID-19 (n=154) | Non-COVID-19 (n=218) |
| --- | --- | --- |
| Accuracy | 96.0% (93.8%-97.8%) |  |
| AUC | 0.986 (0.972-0.996) |  |
| Sensitivity | 92.2% (87.7%-96.2%) | 98.6% (96.9%-100.0%) |
| Specificity | 98.6% (96.9%-100.0%) | 92.2% (87.7%-96.2%) |
| PPV | 97.9% (95.3%-100.0%) | 94.7% (91.6%-97.4%) |
| NPV | 94.7% (91.6%-97.4%) | 97.9% (95.3%-100.0%) |
| F1 score | 95.0% (92.2%-97.4%) | 96.6% (94.8%-98.2%) |

In external validation, COVIDNet significantly outperformed all eight radiologists, which correctly diagnosed 492 of 522 patients, with an accuracy of 94.3% (CI 92.1%-96.1%) and an AUC of 0.981 (CI 0.969-0.990) (Table 2). More impressively, it used only 0.05 minutes on average to diagnose a patient, much faster than the radiologists (1.44 minutes for the fastest radiologist). Cohen’s κ coefficient ^19^ was used to access inter-rater agreement between COVIDNet and the three radiologist groups (all radiologists, the radiologists from the COVID-19 designated hospitals, and the other radiologists) (Supplementary Table 7). Median inter-rater agreement among the radiologists in each group was good, with κ=0.658 [IQR 0.539-0.776], κ=0.738 [IQR 0.680-0.797], and κ=0.600 [IQR 0.502-0.699], respectively. The agreement was good between each group of radiologists and our model (κ=0.745, 0.746, and 0.678, respectively). The agreement between the COVID-19 radiologists and the non-COVID-19 radiologists was excellent (κ=0.853).

**Table 2.** Diagnostic performance and reading time for each patient among COVIDNet and eight radiologists on the external validation cohort. PPV=positive predictive value. NPV=negative predictive value.

|  |  | Accuracy | Sensitivity | Specificity | PPV | NPV | F1-score | Time for each patient (minutes) |
| --- | --- | --- | --- | --- | --- | --- | --- | --- |
|  | COVIDNet | **94.3%**<br>(92.1%-96.1%) | **93.1%**<br>(89.7%-96.2%) | **95.1%**<br>(92.5%-97.5%) | **93.9%**<br>(90.6%-96.8%) | **94.5%**<br>(91.7%-96.9%) | **93.5%**<br>(91.1%-95.7%) | **0.05** |
| COVID-19 radiologists* | Radiologist A (6 yr) | 82.1% | 76.4% | 86.8% | 82.4% | 82.0% | 79.2% | 1.44 |
|  | Radiologist B (8 yr) | 85.1% | 80.7% | 88.6% | 85.1% | 85.1% | 82.8% | 3.66 |
|  | Radiologist C (13 yr) | 83.3% | 73.8% | 91.0% | 86.8% | 81.2% | 79.7% | 4.83 |
|  | Radiologist D (18 yr) | 85.2% | 75.1% | 93.4% | 90.2% | 82.3% | 81.9% | 3.66 |
|  | Average | 83.9% | 76.5% | 90.0% | 86.1% | 79.7% | 80.9% | 3.38 |
| non-COVID-19 radiologists* | Radiologist E (10 yr) | 78.4% | 82.4% | 75.1% | 72.7% | 84.1% | 77.2% | 4.41 |
|  | Radiologist F (11 yr) | 81.0% | 64.8% | 94.1% | 89.9% | 76.9% | 75.3% | 4.83 |
|  | Radiologist G (21 yr) | 72.4% | 45.5% | 94.1% | 86.2% | 68.2% | 59.5% | 4.83 |
|  | Radiologist H (23 yr) | 83.9% | 79.9% | 87.2% | 83.4% | 84.3% | 81.6% | 6.00 |
|  | Average | 78.9% | 68.2% | 87.6% | 83.1% | 78.4% | 73.4% | 5.03 |

The t-SNE representation of chest CT showed two clear clusters, color-coded by the class labels (Figure 2). Most cases located within their respective clusters, suggesting COVIDNet successfully extracted distinct CT features of COVID-19 pneumonia. We selected three groups of representative cases (G1, G2, G3 in Figure 2), and presented their CT manifestations along with probability of COVID-19 in Extended Data Table 8. Typical manifestation of COVID-19 pneumonia is multiple ground glass opacity (GGO) in the subpleural area of bilateral lungs. Radiologists confirmed similar manifestation in the COVID-19 cluster (for example, the G1 red points). As for the misclassified COVID-19 cases (the G3 red points), three cases had no definite finding, which were difficult to identify based the images only; the other cases had extensive GGO with partial consolidation, or combined with pleural effusion and interstitial edema, which were not the typical manifestations of COVID-19. The misclassified non-COVID-19 cases (the G2 blue points) consisted of one bacterial and two influenza B patients, which were classified as COVID-19 due to the appearance of extensive GGO.

**Figure 2.**
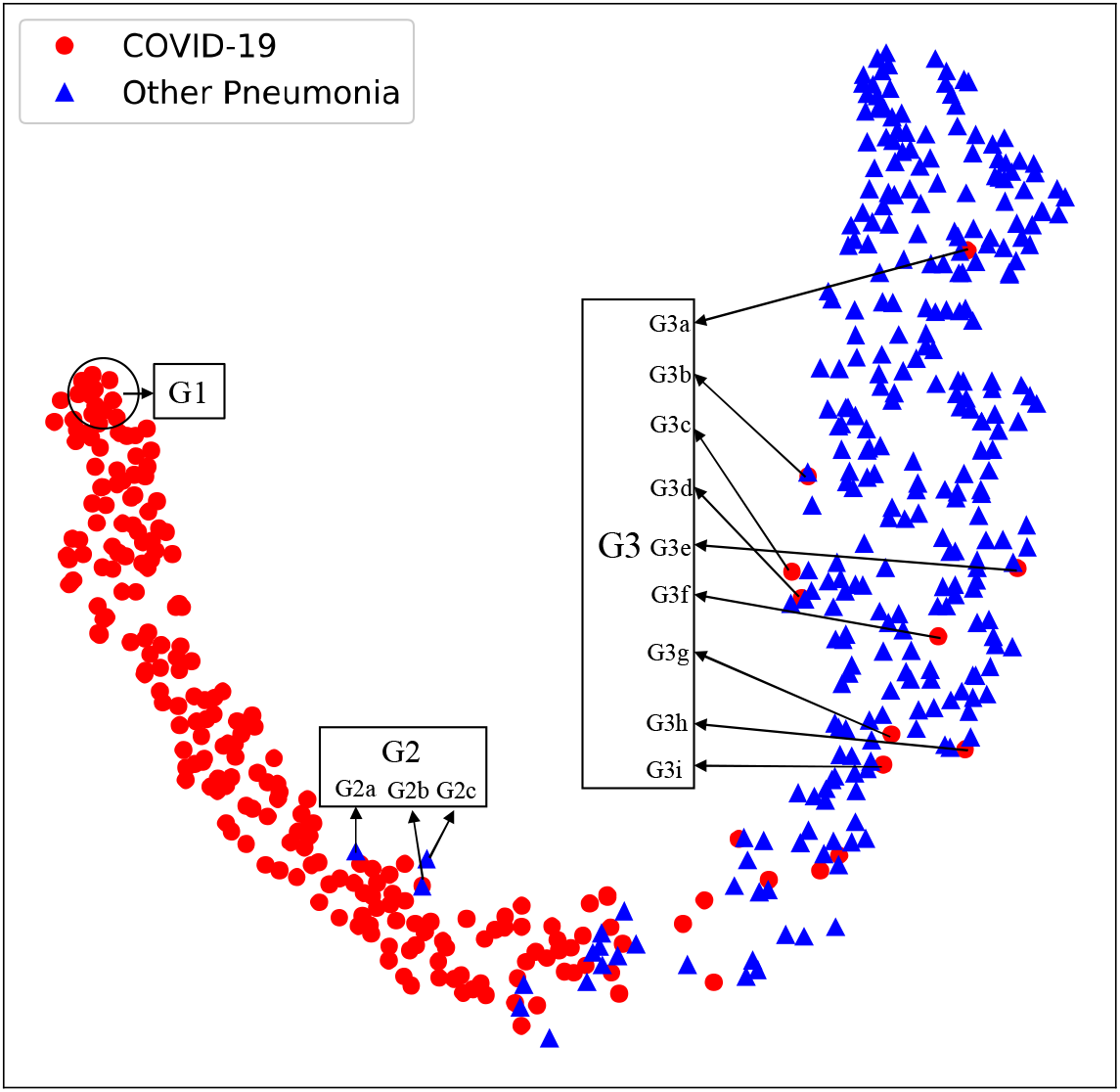
t-SNE of CT images of COVID-19 and other causes of pneumonia on the external test dataset. G1 for chest CT images of COVID-19 pneumonia with highest differentiation. G2 for three cases of false positive prediction of COVIDNet. G3 for nine cases of false negative prediction of COVIDNet.

As of March 23, 2020, COVIDNet has been deployed in 104 hospitals in China, and has helped processing 23,586 CT scans and identification of 5,952 COVID-19 patients. The application pipeline of COVIDNet was illustrated in Extended Figure 3 and 4. The pipeline aided efficiency improvement of the clinical pathway, which offered generalizable clinical insights for regions under considerable strains of nucleic test kits, with limited testing facilities, or facing community transmission epidemic.

Accurate diagnosis of COVID-19 infection is essential for patient management. The specified criteria described in the current COVID-19 clinical management guideline had faced several challenges.^20^ Early clinical manifestations of COVID-19 are fever, cough, and dyspnea that are similar to non COVID-19 viral pneumonia. Chest CT has been a vital part of the COVID-19 infection diagnostic pathway. COVID-19 infection with main CT presentation of GGO can be easily confused with other viral pneumonia and fungal pneumonia. COVID-19 infection with main CT manifestation of consolidation may be confused with bacterial infection.

Our research showed that COVIDNet offered one powerful tool. It could distinguish COVID-19 from other pneumonia infections promptly and accurately. External validation showed COVIDNet’s robustness against seven other types of pneumonia with confirmed pathogen evidence and various CT devices, as well as its faster and more accurate performance over expert radiologists. Our results also showed that the radiologists from COVID-19 designated hospitals performed better than those from the non-epidemic regions. The inter-rater reliability among the radiologists was excellent, combined with their overall poorer performance against COVIDNet, suggested that COVIDNet provided more unbiased results and captured clinically important features of COVID-19 infections that might not have been detected by the human experts, given the fact that all COVID-19 cases were confirmed via nucleic test.

One recent study developed a deep learning screening tool specific to COVID-19.^18^ They extracted features from each axial CT scan of a patient independently, and aggregated the stack of features right before making the classification decision. On the contrary, our COVIDNet model directly extracted spatial features of the entire 3D CT scan using a true three-dimensional deep learning model designed to screen COVID-19 patients. In addition, we demonstrated external generalization ability of our model on an independent external validation dataset through comparison with expert radiologists. Most important of all, COVIDNet has been tested in 104 hospitals of China, and identified 5,962 COVID-19 patients as of March 23^rd^, 2020.

Lacking methods for visualizing how deep learning works has been one of the major bottlenecks for its application in medical settings. To further investigate how the model made classification decisions, we visualized the extracted features using t-Distributed Stochastic Neighbor Embedding (t-SNE).^21^ The results showed that COVIDNet indeed extracted image features that could separate COVID-19 from other types of pneumonia. We also reported image signatures from representatives of the correctly classified COVID-19 cases, the misclassified COVID-19 cases, and the misclassified non-COVID-19 cases. Such image signatures could offer useful insights for clinical decisions. It was an *ad-hoc* analysis of the individuals with interesting allocations in the visualization plot, and the underlying logic and mechanisms of the algorithm still remained unclear. In addition, we could not exclude the possibility that the observed patterns subjected to the patient populations recruited in the external validation study.

When facing an outbreak of COVID-19, often with severe shortage of medical personnel, prompt and accurate image review and interpretation might be a key limiting factor for appropriate clinical decision making. COVIDNet can rapidly detect clinically relevant lung lesions from hundreds of CT images. Together with the probability sorting, COVIDNet may greatly improve the screening and diagnosis efficiency. In addition, COVIDNet is able to automatically quantify the proportion of image abnormalities, supporting further clinical decisions. However, CT presentations for patients with COVID-19 vary dramatically according to stages of the disease, especially for those with basic diseases and complications. In addition, other types of pneumonia may share image abnormalities with COVID-19. Deep learning technology may not perform well under these circumstances. Therefore, patient’s epidemiological and clinical information needs to be closely integrated for further clinical decision making in diagnosis and treatment of COVID-19. The scores produced by the model are not calibrated. Therefore, even though they can serve as a proxy for the classification confidence, their interpretation is rooted in accumulated experience obtained by the integration of model with clinical practice. This virus is evolving in directions which we don’t know yet.^22^ COVIDNet can serve as a cost-effective standing tool for routine screening in clinical settings where chest CT is prescribed.

In conclusion, we have developed an automated classification neural network model, COVIDNet, specifically designed to distinguish COVID-19 from other seven types of pneumonia with confirmed pathogens through analyzing patients’ 3D chest CT scans. In principle, the model can be deployed anywhere in the world with CT imaging capability at a low cost and provide radiological decision support where COVID-19 imaging diagnosis expertise is scarce, especially when facing COVID-19 outbreaks. Our results warrant further validation in future studies.

## Methods

### Datasets

We retrospectively recruited two independent cohorts for model development and external validation with totally 2,800 patients (1,430 COVID-19 patients and 1,370 non-COVID-19 patients) in this study. The model development dataset consisted of CT scans from 2,278 pneumonia patients, who suffered from either COVID-19 or other types of pneumonia. We collected 1,197 COVID-19 cases between January 5, 2020 and March 1, 2020 from ten designated COVID-19 hospitals in China. These COVID-19 cases were confirmed by positive results from RT-PCR assays testing nasal or pharyngeal swab specimens. We also randomly selected 1,081 non-COVID-19 patients with chest CT abnormalities according to the criteria listed in Table S1 from patients that were hospitalized between November 18, 2010 and February 21, 2020 in three other general hospitals in China. CT scan was performed with patients in supine position at full inspiration. All CT scans covered the whole chest.

We excluded 285 patients under the following two circumstances (Figure 1): 244 COVID-19 patients with time intervals between the CT scan and the first positive nucleic acid test longer than two weeks; and 41 patients with severe artifacts, including 33 COVID-19 patients and 8 non-COVID-19 patients.

### Study Ethics

This study was approved by the Ethics Committee (EC) of each participating hospital. The EC has waived the written informed consent requirement for this retrospective study, since the data under evaluation has been de-identified and the research poses no potential risk to patients. The study follows the Declaration of Helsinki. The Trial Registration Number is ChiCTR2000030390 in Chinese Clinical Trail Registry, http://www.chictr.org.cn/showproj.aspx?proj=50224.

### CT Image Collection and Preprocessing

CT images were obtained from different scanners of multiple imaging centers. The detailed CT scan setting and device distribution were listed in Supplementary Table 9 and Extended Figure 5. COVIDNet was a classification neural network that classifies 3D CT images to either the COVID-19 pneumonia class or the non-COVID-19 pneumonia class. CT scans from different sources had various numbers of slices and slice dimensions. For unification, each of the 3D CT scan volume was preprocessed in the following way. We first removed extreme voxel intensities by clipping those outside the range [-1024, 1024] to the interval edges, and linearly scaled the clipped intensities to [0,1]. We subsequently resized the 3D scans to a stack of 64 square axial images of dimension 512 through linear interpolation, and cropped the central square region of size 384 on each 2D axial slice, yielding a stack of 64 axial images of size 384, which was the input to the model. Preprocessed images examples were included in Extended Figure 6.

### Architecture of COVIDNet

The structure of COVIDNet is illustrated in Extended Figure 7, which is a modified DenseNet-264 model consisting of 4 dense blocks^23^. Each dense block has various numbers of composition units. Each unit consists of two sequentially connected stacks with an instance normalization layer^24^, a ReLU activation layer, and a convolution layer. It receives feature maps from all preceding units in the same dense block through dense connections. The batch size in training is 8. We adopted Adam optimizer^25^ with learning rate of 0.001 to minimize the binary cross entropy loss. The model was developed using TensorFlow (version 1.8 with CUDA V9.1.85 and cuDNN 7.0.5) on 16 Tesla P100 GPU.

### Model Evaluation

Accuracy, sensitivity, specificity, positive predictive value (PPV), negative predictive value (NPV), and F1 score were applied to evaluate the performance of the model. Receiver operating characteristic (ROC) curve and confusion matrix were generated based on the classification results. Area under the ROC (AUC) was also calculated. Bootstrap with 10,000 replications was used to calculate the 95% confidence interval of each metric. Evaluation results were obtained and visualized using python libraries, including NumPy, pandas, scikit-learn and Matplotlib.

Furthermore, the performance of COVIDNet was compared with eight independent expert radiologists with 6 to 23 years of experience, on diagnosis of COVID-19 using the external test set. Four radiologists are from the COVID-19 designated hospitals and the other four are not. One research assistant, blinded to the study design, used a stopwatch to record the time that each radiologist spent. In order to ensure the radiologists were spirited, each of them reads CT images up to two hours per day. Before the radiologists initiated the CT image reading, the research assistant informed the CT signs in the guidelines to each radiologist in order to eliminate knowledge bias. The true pneumonia class was blinded to all radiologists. In addition, we used Cohen’s coefficient to evaluate the inter-rater agreement among COVIDNet and the eight radiologists (Table 3).^19^ We categorized κ coefficients as follows: poor (0<κ≤0.20), fair (0.20<κ≤0.40), moderate (0.40<κ≤0.60), good (0.60<κ≤0.80), and excellent (0.80<κ≤1.00).

To further understand the model’s classification decision, we visualized the extracted feature distribution of the model using t-Distributed Stochastic Neighbor Embedding (t-SNE),^21^ which is an unsupervised non-linear dimension reduction algorithm commonly used to visualize high dimensional data. It projects high dimensional feature maps right before the final fully connected layer of COVIDNet onto a two-dimensional space, and converts similarities between the original data pairs to similarities between the projected data pairs in the two-dimensional space. Since it considers the local structure so that after projection, it can reveal interesting clusters among the data points.

### Code and Data Availability

The code and CT image data, as well as clinical data were available only for research after obtaining the agreement of corresponding authors.

## Acknowledgments

This study was supported by National key research and development program [2017YFC0114001]. COVIDNet was developed in collaboration with Biomind (https://biomind.ai), which played an important role in code programming.

## Author Contributions

Kunlun He, Jianxin Zhou, Hao Li, Haipeng Shen had roles in the study design. Peng Xie,Yiqing Tan, Jicheng Du, Zhibing Zhang, Xia Fu, Yanqing Fan, Meifang Li, Na Zhang, Lin Li, Yaou Liu, Lin Xu, Jingbo Du, Zhenhua Zhao, Xuelong Hu, Jianmin Cheng, Weipeng Fan, Rongpin Wang, Weimin An is responsible for data collection. Gao Huang and Pan Liu designed the model. Gao Huang, Minchao Liu and Huayuan Guo consulted on the code. Pan Liu and Zongren Li programmed the model. Bohan Liu, Lutao Dai, Yanlin Yang, Qin Zhong, Yuhui Sun and Wenjun Wang worked on statistical aspects of the study. Xizhou Guan, Ning Xing, Chongchong Wu, Yongkang Nie, Liuquan Cheng, Lin Ma and Weimin An has roles in data interpretation, Bohan Liu, Lutao Dai, Chenghui Zhao, Wei Shan and Hao Li take responsibility for the integrity of the data and the accuracy of the data analysis. Bohan Liu, Pan Liu, Lutao Dai and Chenghui Zhao made the figures. Qin Zhong, Xixiang Lin, Yuhui Sun, Zongren Li and Qian Jia did literature search. All authors approved the final version for submission.

## Competing interests

We declare no competing interests.

## Notes

### Competing Interest Statement

The authors have declared no competing interest.

### Clinical Trial

ChiCTR2000030390

### Clinical Protocols

http://www.chictr.org.cn/showproj.aspx?proj=50224

